# Real-World Evidence on Dose-Reduction and Treatment Outcomes in Small Cell Lung Cancer; A Bayesian mixed effects and Competitive Risk Approach

**DOI:** 10.1101/2024.12.04.24318490

**Authors:** Luca Marzano, Asaf Dan, Adam S. Darwich, Luigi De Petris, Salomon Tendler, Rolf Lewensohn, Jayanth Raghothama, Sebastiaan Meijer

## Abstract

**Background:** Small cell lung cancer (SCLC) is a challenging disease to treat due to rapid progression, development of chemo-resistance, and discrepancies in outcomes between real-world data and clinical trials. Previous studies lack comprehensive analyses of intermediate events and the treatment process, such as treatment decisions, progression of disease, and the occurrence of adverse events (AEs) over time. The aim of this study was to apply advanced statistical methods on a longitudinal SCLC data set in order to identify factors of importance for the risk of AEs and for survival.

**Methods:** Information was collected on the treatment pathways of 421 SCLC patients treated at the Karolinska University Hospital between 2016-2022 (Stockholm, Sweden). Analysis focused on the impact of dose-adjustment on adverse events (AEs), including neutropenia, by estimating odds ratios (OR) using Bayesian mixed-effects modelling. Covariate’s effects on ECOG performance status (PS) deterioration and early discontinuation of chemotherapy with cause-specific hazard ratios (csHR) were explored using competitive risk models. This approach was applied to cohorts of patients receiving first line platinum etoposide, and second line platinum etoposide or platinum irinotecan.

**Results:** At the end of the first line treatment, most patients exhibited tumour regression (n=167). Patients with neutropenia had longer overall survival (HR: 0.70 [0.53, 0.92]). Higher etoposide dose levels were associated with subsequent occurrences of AEs (OR: 5.97 [1.41, 30.5]) and neutropenia (OR: 3.55 [1.03, 13.3]). Dose adjustment did not affect overall survival as long as the patient completed the four-dose regimen treatment. For the second-line, fewer patients completed four treatment cycles and the most common reason of early discontinuation was tumour progression (n=72). Male patients experienced fewer AEs and better first line treatment response compared to females (csHR: 0.51 [0.25, 0.90]). High-risk patients (here defined as ECOG PS 2-3, or age over 75 years) with early discontinuation of therapy had survival outcomes similar to those who did not receive therapy.

**Conclusions:** Our results indicate that SCLC therapies may benefit from more individualized dosing strategies. These strategies would aim to balance improved survival with reduced risk of AEs, particularly neutropenia. It would also be beneficial to assess the risk-benefit of treating specific subgroups, including patients receiving second line therapy. Real-world data are crucial for studying therapy response and risk-benefit of treating patient groups that are underrepresented in clinical trials.

## Introduction

Small cell lung cancer (SCLC) is an aggressive and challenging disease to treat, mainly due to the lack of knowledge surrounding chemo-resistance mechanisms^1,2^. Standard SCLC treatment consists of defined treatment lines, each typically comprising four cycles of platinum-based chemotherapy (Cisplatin or Carboplatin) with Etoposide (PE) or Irinotecan (IP), aimed at achieving initial treatment effects, followed by a planned rest period of 2–3 months. For patients with cancer stages I–III, this is often combined with concurrent thoracic radiotherapy (RT). In stage IV cancer, immunotherapy with checkpoint inhibitors can be combined with chemotherapy, though with modest survival benefits^3,4^. However, most SCLC patients experience relapse within 6 months after initial treatment, necessitating further therapy, though with limited success^5^.

In the setting of disease relapse, the distinction between sensitive- and resistance-relapse plays a crucial role in predicting survival. Patients with sensitive disease, defined by relapse occurring more than 90 days after the completion of first-line platinum-based chemotherapy, might be re-challenged with the same chemotherapy regimen administered as first-line, and demonstrate significantly better survival outcomes compared to those with resistance disease, where relapse occurs within 90 days^6,7^. In this latter case, the most common approach is to change to other chemotherapy agents, if the patient is still in acceptable general condition.

The rapid progression of SCLC often makes it difficult to study and manage^8^ which results in discrepancies in observed outcomes between clinical trials and real-world patients^9^. The knowledge gap between clinical trials and the real-world patient population is overcome through treatment adjustments based on clinical experience, including choice of chemotherapy agents, dose adjustments, and end-of-life care^8,10,11^. The focus of previous studies has mainly been on survival outcomes, often overlooking patients who received subsequent treatment lines after first-line chemotherapy. Further, there is a lack of studies that analyse events and intermediary events that occur over time in the treatment process^8,12–14^.

Real-world data (RWD) offers an opportunity to retrospectively explore the impact of treatment decisions and pathways on patient outcomes, given the variability in the real-world patient population^15,16^. As such, RWD has the potential to inform the individualisation of treatments for specific subpopulations^17,18^, such as patients with poor ECOG (Eastern Cooperative Oncology Group) performance status (PS), older persons, and patients with brain metastases who tend to be underrepresented in clinical trials^9^.

However, evidence generation from RWD is challenging because of complex clinical workflows and issues related to its secondary use, such as inherent biases in data collection and decision-making processes, and non-observable effects^18,19^. In SCLC treatment, the nuances of dose adjustments, planned treatment pauses, assessment of response, and early discontinuation of treatment due to severe toxicity or health deterioration, exemplify the intricacies involved^20,21^.

The aim of this study is to describe the time-dynamics of patient trajectories in a clinical SCLC patient cohort, in order to identify factors of importance for dose-adjustments and survival outcomes. This approach necessitates an appropriate choice of data analysis^22^. While artificial intelligence and machine learning have shown much promise for analysing RWD, issues of transparency and interpretability pose a potential risk of propagating biases from the data^23^. Here the focus was on selecting transparent modelling approaches^23,24^, allowing for clinical contextualisation and interpretation^9,25^.

The analysis consisted of a combination of Bayesian^26–28^ and mixed-effects models^10,29,30^ for studying dose adjustments and their impact on AEs (Bayesian logistic regression^31^) and the grade of toxicity (Bayesian cumulative link ordinal regression^31^). Competitive risk Fine-Gray regression^32^ was applied to compute cause-specific hazard ratios and predict the time-course of the therapy, ECOG PS deterioration and early discontinuation of treatment, defined as discontinuation before completing four planned chemotherapy cycles due to AEs, or radiological and clinical signs of tumour progression.

## Materials and Methods

### Collection of longitudinal data

The study received ethical approval by the Swedish Ethical Review Authority (approval number: 2023-07389-01). Patients still alive at the time of data collection signed a written informed consent. The study included a total of 421 patients diagnosed with SCLC at the Karolinska University Hospital in Stockholm, Sweden, between 2016 and 2022. Baseline patient characteristics such as age, sex, smoking status, TNM (Tumour, Node, Metastasis) stage (IASLC, International Association for the Study of Lung Cancer, 8th edition), Charlson Comorbidity Index (CCI), CNS metastasis, and previous cancer prior to SCLC were recorded. ECOG PS was recorded at baseline and at the start and cessation of each treatment line. Blood values, including haemoglobin, creatinine, sodium (Na), lactate dehydrogenase (LDH), C-reactive Protein (CRP), and albumin, were computed at baseline and at the start of each treatment line.

The specific drug regimen received by each patient was recorded. Body surface area (BSA) and estimated glomerular filtration rate (GFR) were recorded before start of treatments. These two variables were used to compute the dose of chemotherapy administered for each patient (e.g., GFR for carboplatin dosing using Calvert formula^33^, and BSA for etoposide and irinotecan, See Supplementary Table S1). The date and percentage of dose administered for each chemotherapy treatment were registered, including the reason for any modifications or adjustment to the administrated doses, for the calculation of dose interval and intensity.

The reason for dose reduction was recorded. Adverse events (AEs) were classified by type and grade of severity of toxicity (GT) according to the Common Terminology Criteria for AEs (CTCAE) version 5. In this work, high toxicity was defined as AEs with CTCAE GT 3-4.

Details on radiotherapy, including the treatment indication (curative, consolidating, as a separate treatment line or palliative) and fractionation were reported. At the end of each treatment line, the best response achieved was scored as Complete or Partial Response (CR or PR, respectively), Stable Disease (SD) or Progressive Disease (PD).

The overall survival (OS) was calculated from the date of diagnosis until the patient’s death or the end of the study period, whichever came first. For each treatment line, Progression-free survival (PFS) was calculated from the date of the administration of the first dose, until signs of progression or death.

### Analysis description

Several aspects of the treatment process were analysed for each treatment line, including: dose-adjustments due to AEs, changes in ECOG PS from the start to the end of the treatment line, and early treatment discontinuation (stopping treatment before 4 cycles of chemotherapy due to signs of tumour progression, toxicity, deteriorating medical condition, or unwillingness of the patient to receive the treatment) versus no – early discontinuation, defined as cessation of a treatment after 4 cycles of chemotherapy, followed by a pause due to PR or SD.

The analytical pipeline can be summarized in the following steps:

- selection of clinical scenarios and patient subgroups
- exploratory data analysis of the longitudinal cohort
- multivariate analysis of dose adjustments
- multivariate analysis of competitive risks
- analysis of high risk subgroups

All analyses and plotting of results were carried out using RStudio, version 2022.12.0 + 353.

### Selection of clinical scenarios and patient subgroups

Prior to commencing the analysis, the collected data was analysed with a broad perspective to identify key subpopulations of interest. The cohort was explored in terms of baseline characteristics at the first line treatment decision, and the administered chemotherapy lines using summary statistics and Sankey Flows for the subsequent treatment lines. Treatment lines with relevant sample size were detected for the next analysis steps. Moreover, special subgroups of patients receiving treatment but with high fragile conditions were selected.

### Exploratory analysis of the longitudinal cohort

The effects of AEs, namely CTCAE G3-4 AEs, ECOG PS change at the end of treatment, and early discontinuation of therapy were assessed by exploring absolute count and relative percentages. Hazard ratios (HRs) and Kaplan Meier curves were computed to estimate the impact on survival outcomes^34^. The cumulative incidence-time curves of dose-reduction due to AEs, separating patients with good therapy response and the ones with unplanned stops, were analysed^35^. Moreover, the survival outcomes were estimated by stratifying patients by those received four doses (full course) without dose reduction, patients that received four cycles but with dose reduction, and patients that received less than four cycles.

### Multivariate analysis of dose adjustments

For the dose adjustment analysis, the association between dose reductions caused by an AE and the grade of toxicity was explored. This was done by training two Bayesian mixed-effect regression models: a binary logistic regression for the adverse events incidence^31,36^, and a cumulative link regression for the grade of toxicity modelled as an ordinal outcome^31,37^. The longitudinal variables (dose level, dose interval, and their mutual interaction) were analysed with respect to the key baseline characteristics. The Bayesian logistic regression can be summarised as follows:

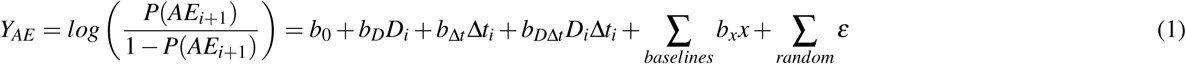

Where *P*(*AE_i_*_+1_) is the probability of a dose reduction due to adverse event at dose *i* + 1 caused by the *i^th^* dose *D_i_*, with interval Δ*t_i_* between the doses, *x* baseline covariates and *ε* random effects. The cumulative link ordinal Bayesian regression can be summarised as follows:

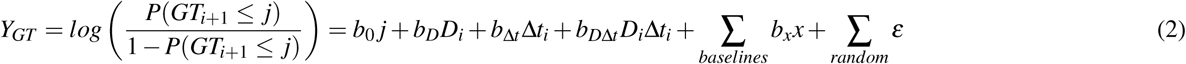

With similar notation for the Bayesian logistic regression, where *P* (*GT_i_*_+1_ *≤ j*) is the probability of grade of toxicity lower than or equal to *j* (with *j* = 0*,…,* 4), and *b*_0_ *_j_* is the baseline intercept for the *j^th^*level. The Bayesian models were trained running four independent Markov chains Monte Carlo (MCMC) with 2,500 iterations for each chain. The odds ratios (exponential of the coefficients b) with the 95% confidence level were estimated for all the covariates. The random effects were estimated for the individual patients, administrated doses and the dose interval. This allowed for individual differences and the time-dependent nature of the dose variables to be considered.

### Multivariate analysis of competitive risks

For the variation of ECOG PS at the end of the treatment line and the early discontinuation of the therapy, a competitive risk time-to-event model was developed. Fine-Gray competitive risk regression was used with the outcomes^31^, the progression free survival and the event of interest to estimate the distributed Hazard ratios of the covariates. The model can be summarised as follows:

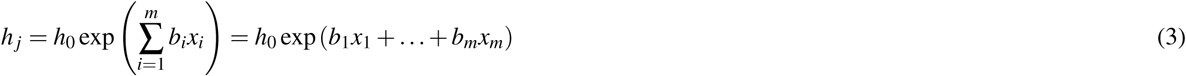

Where, *h_j_* is the cause-specific hazard of the event of interest, *h*_0_ the baseline hazard, and the exponential of the *m^th^* coefficient *b* the distributed hazard ratio of the *m^th^* covariate *x*. The cause-specific hazard ratios were computed for ECOG PS deterioration/improvement (in competition with unchanged PS or improvement/deterioration), and the unplanned stop of the therapy in competition with planned stop of statement due to PR or SD. Here we indicate the OS or PFS Cox hazard ratios with HR, csHR for the cause-specific hazard ratios computed with the Fine-Gray model, and OR the odds ratios obtained by the Bayesian mixed-effects models.

### High-risk subgroups’ analysis

Multivariate analysis was performed for the subgroups of patients with detected fragile conditions receiving treatment. The OS of these patients was compared to the OS of patients with similar characteristics that did not receive chemotherapy. This allowed the comparison of survival outcomes of patients with poor response with the ones who did not receive chemotherapy, thus comparing the benefits and risks of the received treatment.

## Results

### Selection of clinical scenarios and patient subgroups

The patient cohorts taken into consideration were the subjects receiving platinum etoposide (PE) as first line therapy, and the patients receiving either rechallenge with PE, or platinum and irinotecan (IP) as second line therapy. The analysis included stage III-IV patients and excluded patients with ECOG PS of 4 (n=29). Supplementary Figure S1 reports the inclusion/exclusion flowchart of the cohort. In terms of toxicity, only neutropenia was analysed in detail in this study, since it was the most commonly reported treatment-related AE (See Supplementary Table S2). Supplementary Figures S1-2 show Sankey flows of the subsequent treatment lines and the dose adjustments.

The identified high-risk subgroups included the first line patients receiving platinum etoposide with ECOG PS 2-3, patients with CNS metastases, over the age of 75 years, and patients with advanced stage IVB. These subgroups were selected since they are known to be either underrepresented in RCTs or are associated with relatively poor OS compared to the general population^9^. The age threshold was chosen to define a sharp subgroup of elderly patients from the cohort. The multivariate analysis was not performed for patients with CNS metastases due to wide confidence intervals of ORs and csHR caused by the small sample size of the subgroup (n=42).

For some individuals, blood values and lab tests were missing (from which the variables with higher percentage of missing values in the entire cohort were neutrophil counts 44.9%, LDH 22.9%, and CRP 18.3%). The percentages of missing values for neutrophil counts at the start of the treatment lines were as follows: 50.9% in the first line of platinum etoposide, 34.8% in the second line platinum etoposide, and 39.3% in the second line of platinum irinotecan. The high percentage of missing neutrophil values was due to these not being routinely measured at baseline before the initiation of chemotherapy treatment. Therefore, during the multivariate analyses, for each treatment line analysis, values were imputed using the machine learning algorithm MissForest^38^. MissForest was chosen since it is particularly effective for datasets with mixed-type variables and complex interactions, maintaining low imputation errors and preserving relationships among variables even with high percentages of missing information^39^.

### Exploratory survival analysis

In the first line setting, chemotherapy doses were reduced in 127 out of 289 patients (43.94%), mainly due to any grade of neutropenia (n=89) and AE GT 3-4 (n=104) (Table 1). Similar figures were observed for the second line setting (PE n=69, IP n=56), with neutropenia mainly associated with PE treatment (n=26). At the end of the first line, n=179 (out of which n=167 PR) patients did not experience early discontinuation of treatment. Early discontinuation of therapy happened in 110 cases, due to clinical (n=48) or radiological (n=23) signs of PD. In the second line, early discontinuation (n=72) was more common than completion of treatment (n=52).

**Table 1.**
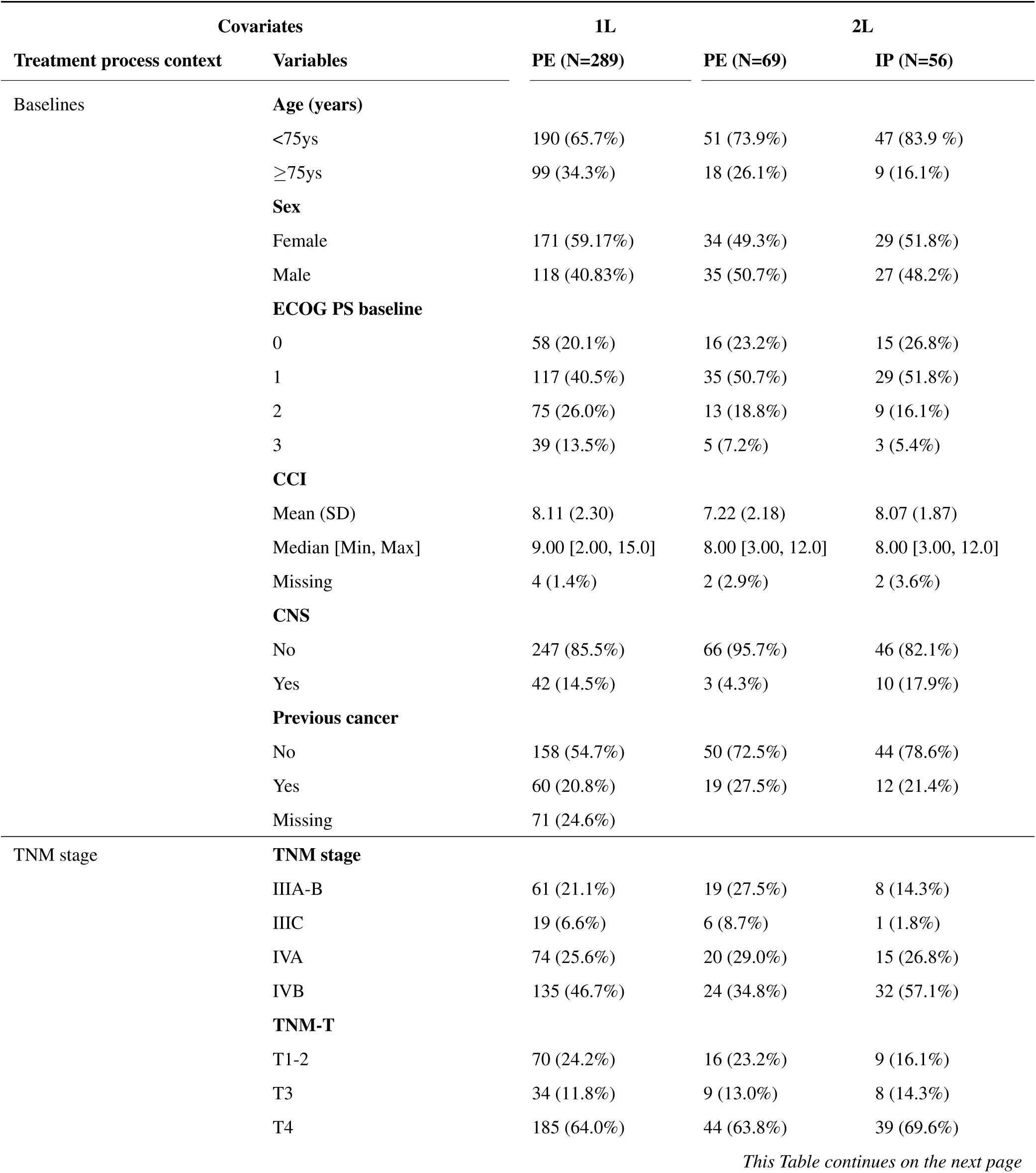
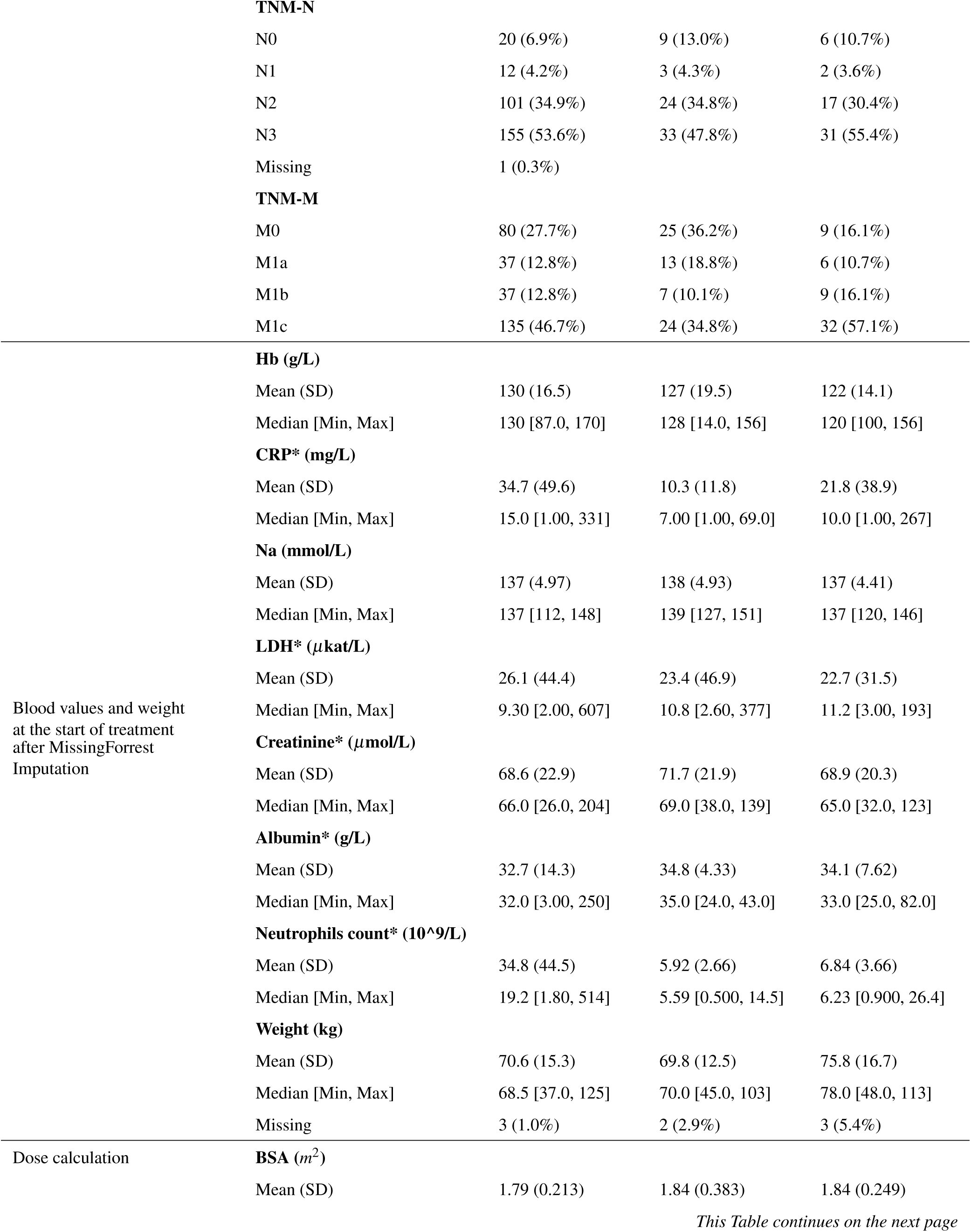
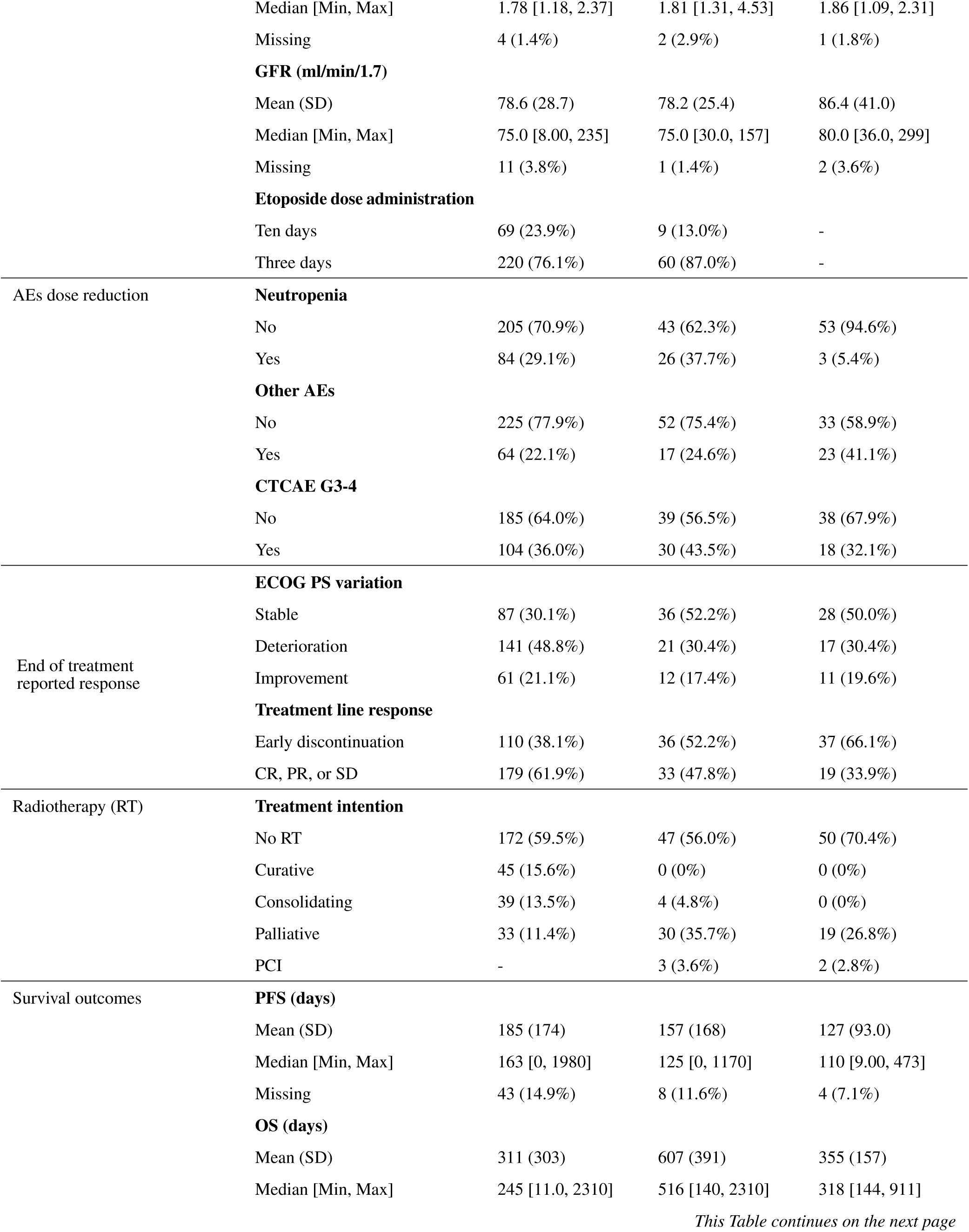
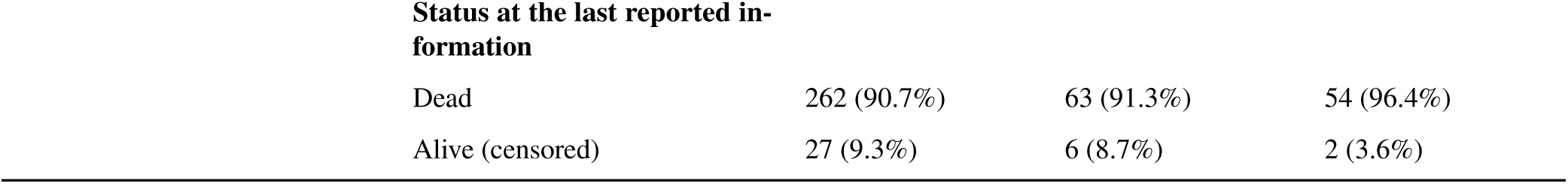
Summary of the analysed cohort. 1L: first line, 2L: second line, PE: platinum etoposide, IP platinum Irinotecan, ECOG PS: performance status, CCI: Charles Comorbidity Index, CNS: brain metastasis, LDH: lactate hydrogenasis, CRP: c-reactive protein, Na: sodium, BSA: body surface area, GFR: estimated glomerular filtration rate, AEs: adverse events, PFS: progression free survival, OS: overall survival. SD: standard deviation. (*): log transformation applied during the analysis to handle high skewness.

Figure 2 shows the cumulative incidences of dose adjustment due to neutropenia. The incidence was higher for patients in both lines who achieved treatment completion without premature discontinuation. This was not the case for other AEs (Figure 2). In the first line scenario, occurrences of neutropenia and other AEs were associated with more favourable OS (HR: 0.70 [0.53, 0.92] and HR: 0.72 [0.57, 0.95] respectively). In the second line, only neutropenia was associated with better OS (HR: 0.51 [0.33, 0.80]). OS of patients completing four cycles of first-line treatment was not affected by dose reduction due to AEs (HR: 0.85 [0.59, 1.23]) (Figure 3).

**Figure 1.**
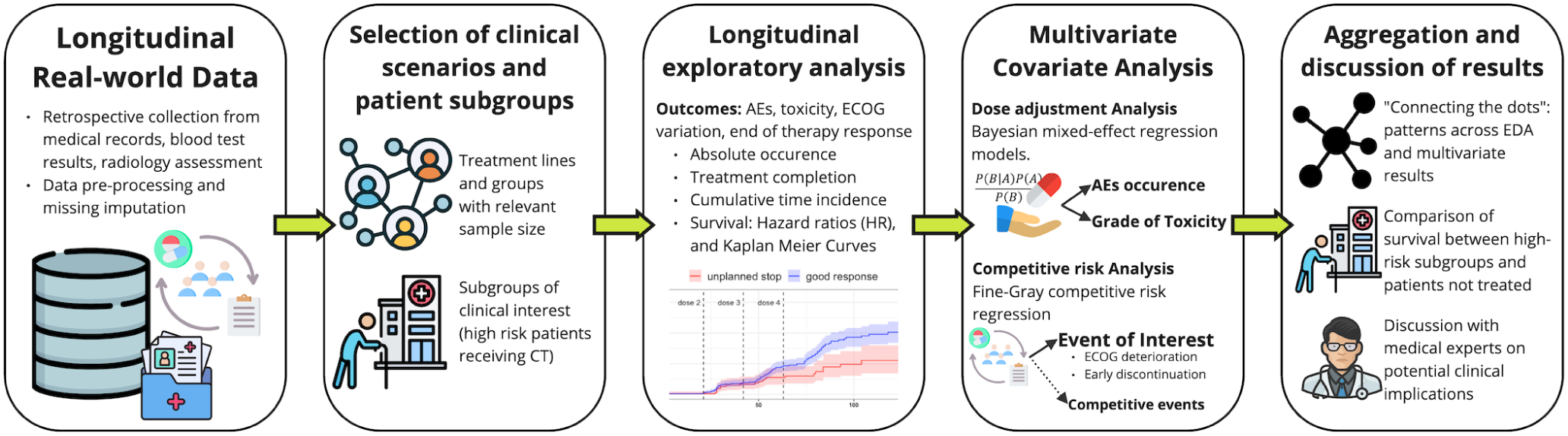
Analytical pipeline description.

**Figure 2.**
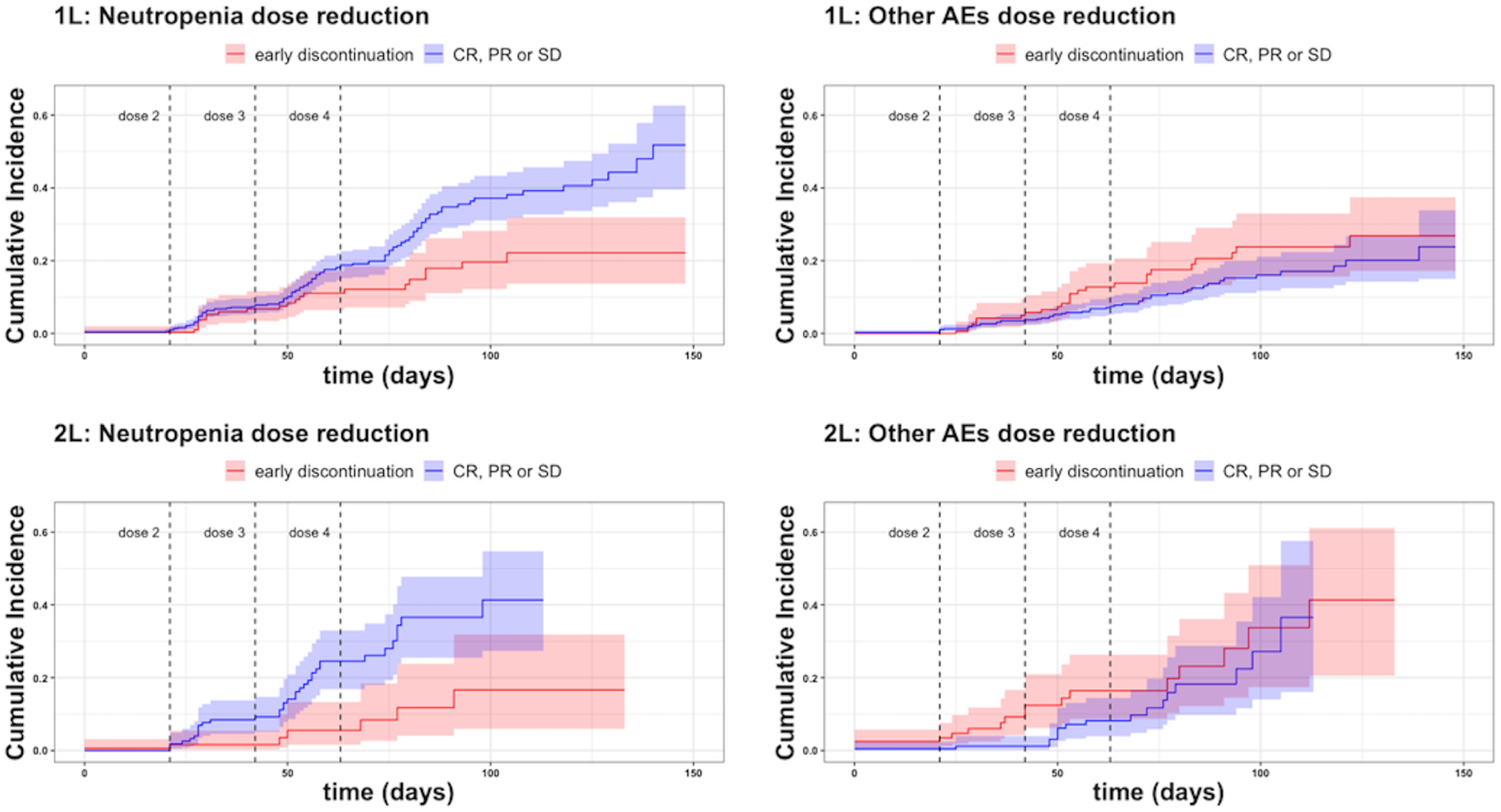
Cumulative incidences of dose reduction (%) due to neutropenia and other adverse events (AEs) for the first and the second line therapy. The curves are stratified by early discontinuation of chemotherapy (red curves) and therapy response (CR complete, PR partial, SD stable disease) (blue curves). Dotted lines correspond to the three week intervals corresponding to planned administration of the second, third and fourth doses.

**Figure 3.**
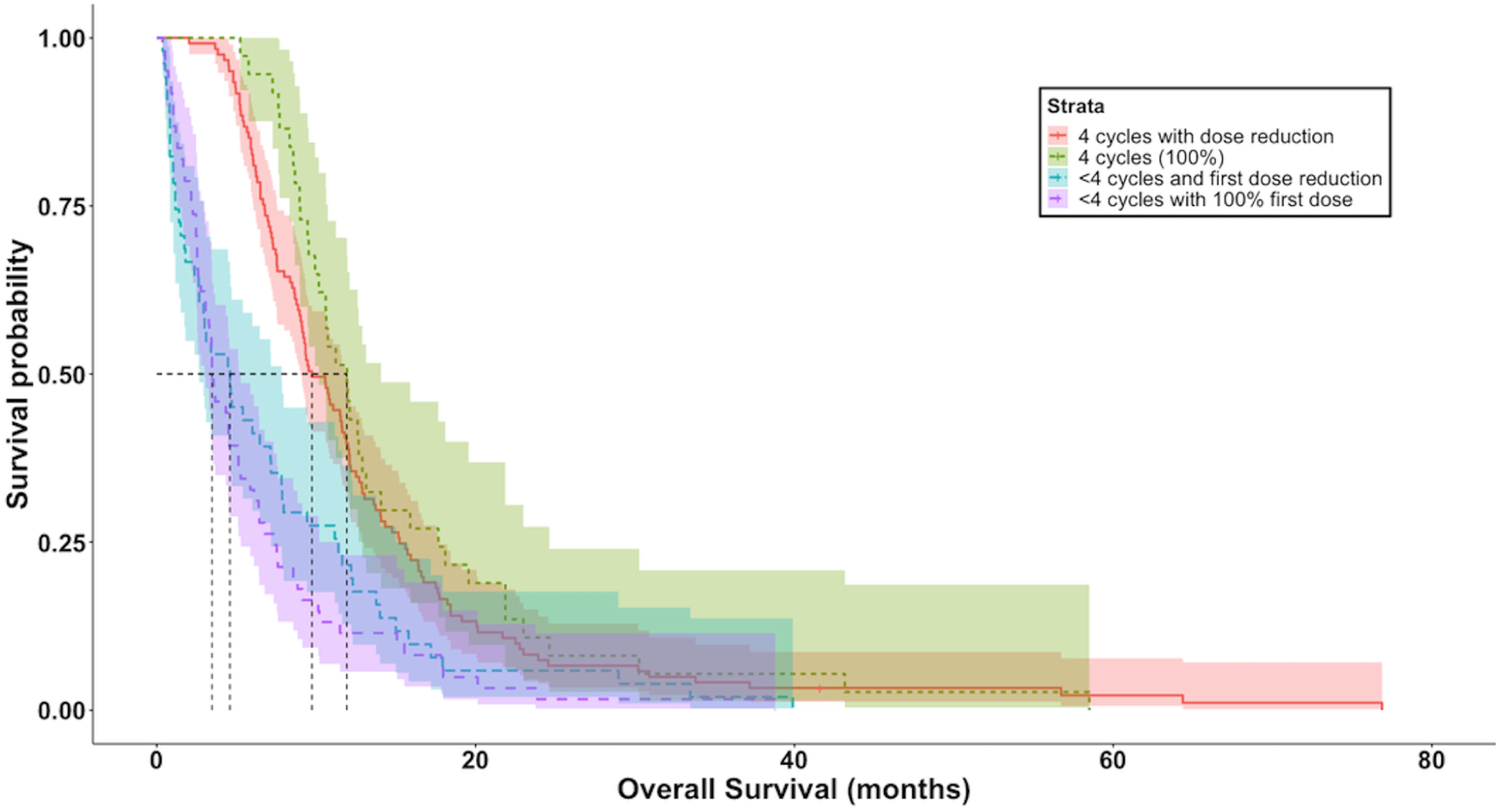
Kaplan Meier curves for overall survival of patients receiving first line treatment, stratified by administrated doses.

Patients with early discontinuation of therapy in the first line setting, showed a markedly worse OS compared to patients who completed 4 cycles chemotherapy (HR: 4.27 [3.27, 5.57]). Similarly, worse OS was observed for patients with early discontinuation during second line therapy (HR: 2.21 [1.50, 3.27]). Moreover, ECOG PS deterioration was significantly associated with a poorer prognosis, even if this was only seen in the first line setting (HR: 2.05 [1.54, 2.72], reference: stable ECOG PS).

Patients receiving PE as second line completed the four-dose cycle more often than the IP patients (73.36% vs 41.07%, respectively). As expected, second line PE patients showed better OS (HR: 0.51 [0.33, 0.78], overall survival computed from the start of the second line). The PFS of PE patients was similar compared to patients receiving IP (HR: 0.84 [0.57, 1.23]).

### Multivariate analysis

#### First line therapy PE

Tables 2 and 3 detail the results of the dose adjustment and the competitive risk analysis for the first line therapy. All the longitudinal variables presented significant variability (See Supplementary Table S3). Inter-individual variability was higher than inter-occasion variability.

**Table 2.**
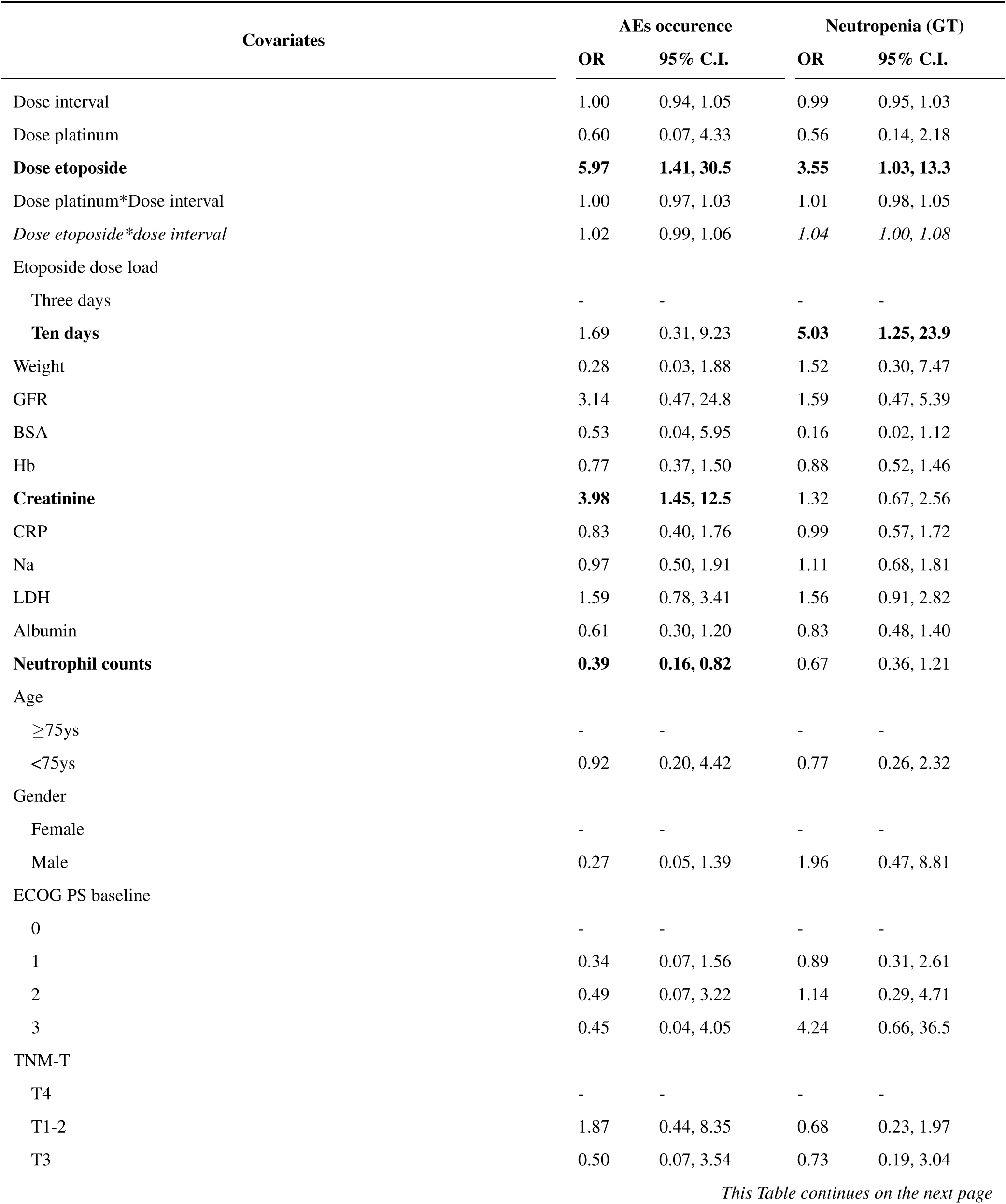
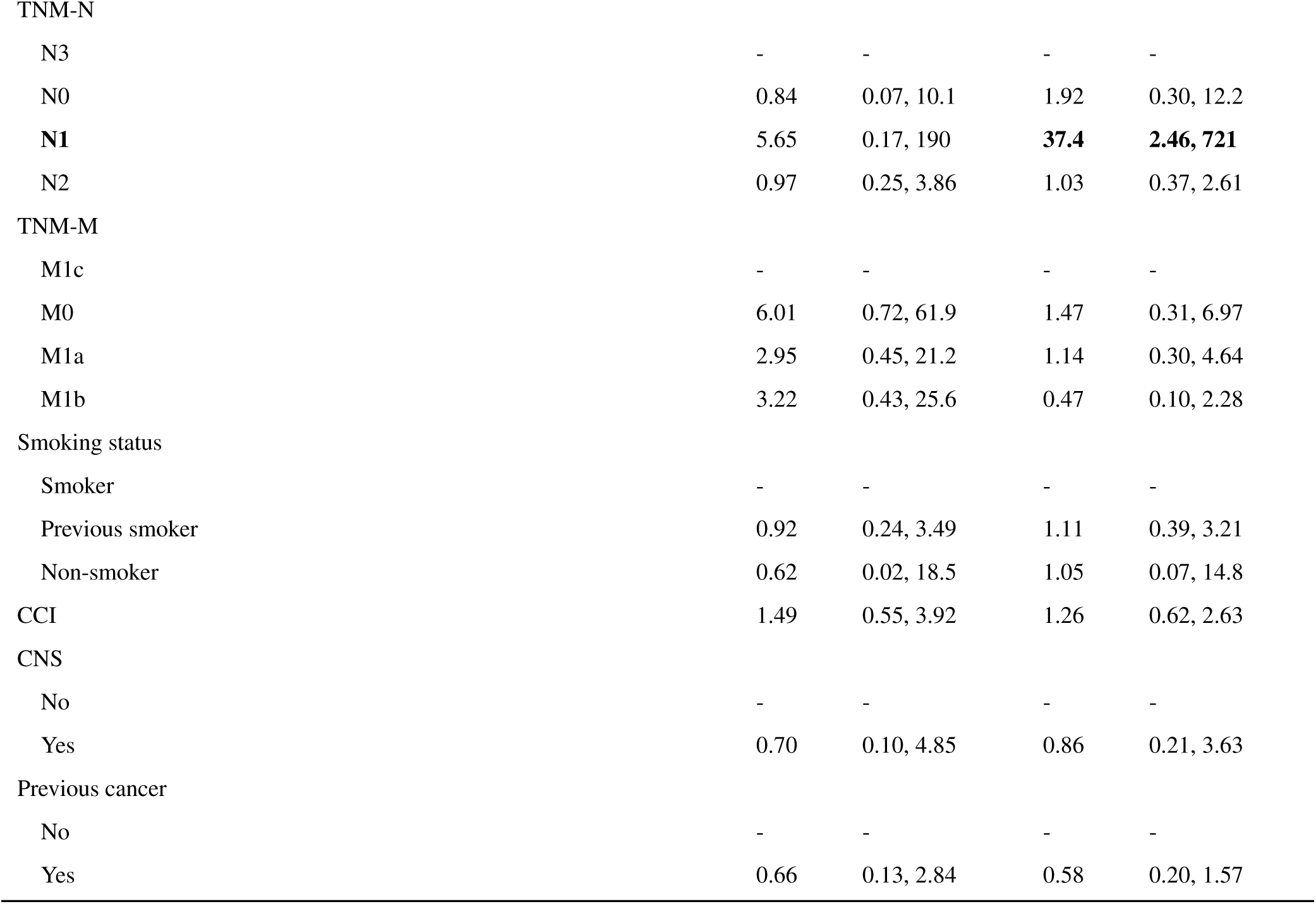
Bayesian mixed-effects analysis for the first line platinum etoposide patients. AEs: adverse effects, GT: grade of toxicity, OR: Odds ratio, C.I.: confidence interval, ECOG PS: performance status, CCI: Charles Comorbidity Index, CNS: brain metastasis, LDH: lactate hydrogenasis, CRP: c-reactive protein, Na: sodium, Hb: hemoglobin, BSA: body surface area, GFR: estimated glomerular filtration rate.

| Covariates | AEs occurrence |  | Neutropenia (GT) |  |
| --- | --- | --- | --- | --- |
|  | OR | 95% C.I. | OR | 95% C.I. |
| Dose interval | 1.00 | 0.94, 1.05 | 0.99 | 0.95, 1.03 |
| Dose platinum | 0.60 | 0.07, 4.33 | 0.56 | 0.14, 2.18 |
| **Dose etoposide** | **5.97** | **1.41, 30.5** | **3.55** | **1.03, 13.3** |
| Dose platinum*Dose interval | 1.00 | 0.97, 1.03 | 1.01 | 0.98, 1.05 |
| *Dose etoposide\*dose interval* | 1.02 | 0.99, 1.06 | *1.04* | *1.00, 1.08* |
| Etoposide dose load |  |  |  |  |
| Three days | - | - | - | - |
| **Ten days** | 1.69 | 0.31, 9.23 | **5.03** | **1.25, 23.9** |
| Weight | 0.28 | 0.03, 1.88 | 1.52 | 0.30, 7.47 |
| GFR | 3.14 | 0.47, 24.8 | 1.59 | 0.47, 5.39 |
| BSA | 0.53 | 0.04, 5.95 | 0.16 | 0.02, 1.12 |
| Hb | 0.77 | 0.37, 1.50 | 0.88 | 0.52, 1.46 |
| **Creatinine** | **3.98** | **1.45, 12.5** | 1.32 | 0.67, 2.56 |
| CRP | 0.83 | 0.40, 1.76 | 0.99 | 0.57, 1.72 |
| Na | 0.97 | 0.50, 1.91 | 1.11 | 0.68, 1.81 |
| LDH | 1.59 | 0.78, 3.41 | 1.56 | 0.91, 2.82 |
| Albumin | 0.61 | 0.30, 1.20 | 0.83 | 0.48, 1.40 |
| **Neutrophil counts** | **0.39** | **0.16, 0.82** | 0.67 | 0.36, 1.21 |
| Age |  |  |  |  |
| ≥75ys | - | - | - | - |
| <75ys | 0.92 | 0.20, 4.42 | 0.77 | 0.26, 2.32 |
| Gender |  |  |  |  |
| Female | - | - | - | - |
| Male | 0.27 | 0.05, 1.39 | 1.96 | 0.47, 8.81 |
| ECOG PS baseline |  |  |  |  |
| 0 | - | - | - | - |
| 1 | 0.34 | 0.07, 1.56 | 0.89 | 0.31, 2.61 |
| 2 | 0.49 | 0.07, 3.22 | 1.14 | 0.29, 4.71 |
| 3 | 0.45 | 0.04, 4.05 | 4.24 | 0.66, 36.5 |
| TNM-T |  |  |  |  |
| T4 | - | - | - | - |
| T1-2 | 1.87 | 0.44, 8.35 | 0.68 | 0.23, 1.97 |
| T3 | 0.50 | 0.07, 3.54 | 0.73 | 0.19, 3.04 |
| TNM-N |  |  |  |  |
| N3 | - | - | - | - |
| N0 | 0.84 | 0.07, 10.1 | 1.92 | 0.30, 12.2 |
| **N1** | **5.65** | **0.17, 190** | **37.4** | **2.46, 721** |
| N2 | 0.97 | 0.25, 3.86 | 1.03 | 0.37, 2.61 |
| TNM-M |  |  |  |  |
| M1c | - | - | - | - |
| M0 | 6.01 | 0.72, 61.9 | 1.47 | 0.31, 6.97 |
| M1a | 2.95 | 0.45, 21.2 | 1.14 | 0.30, 4.64 |
| M1b | 3.22 | 0.43, 25.6 | 0.47 | 0.10, 2.28 |
| Smoking status |  |  |  |  |
| Smoker | - | - | - | - |
| Previous smoker | 0.92 | 0.24, 3.49 | 1.11 | 0.39, 3.21 |
| Non-smoker | 0.62 | 0.02, 18.5 | 1.05 | 0.07, 14.8 |
| CCI | 1.49 | 0.55, 3.92 | 1.26 | 0.62, 2.63 |
| CNS |  |  |  |  |
| No | - | - | - | - |
| Yes | 0.70 | 0.10, 4.85 | 0.86 | 0.21, 3.63 |
| Previous cancer |  |  |  |  |
| No | - | - | - | - |
| Yes | 0.66 | 0.13, 2.84 | 0.58 | 0.20, 1.57 |

**Table 3.**
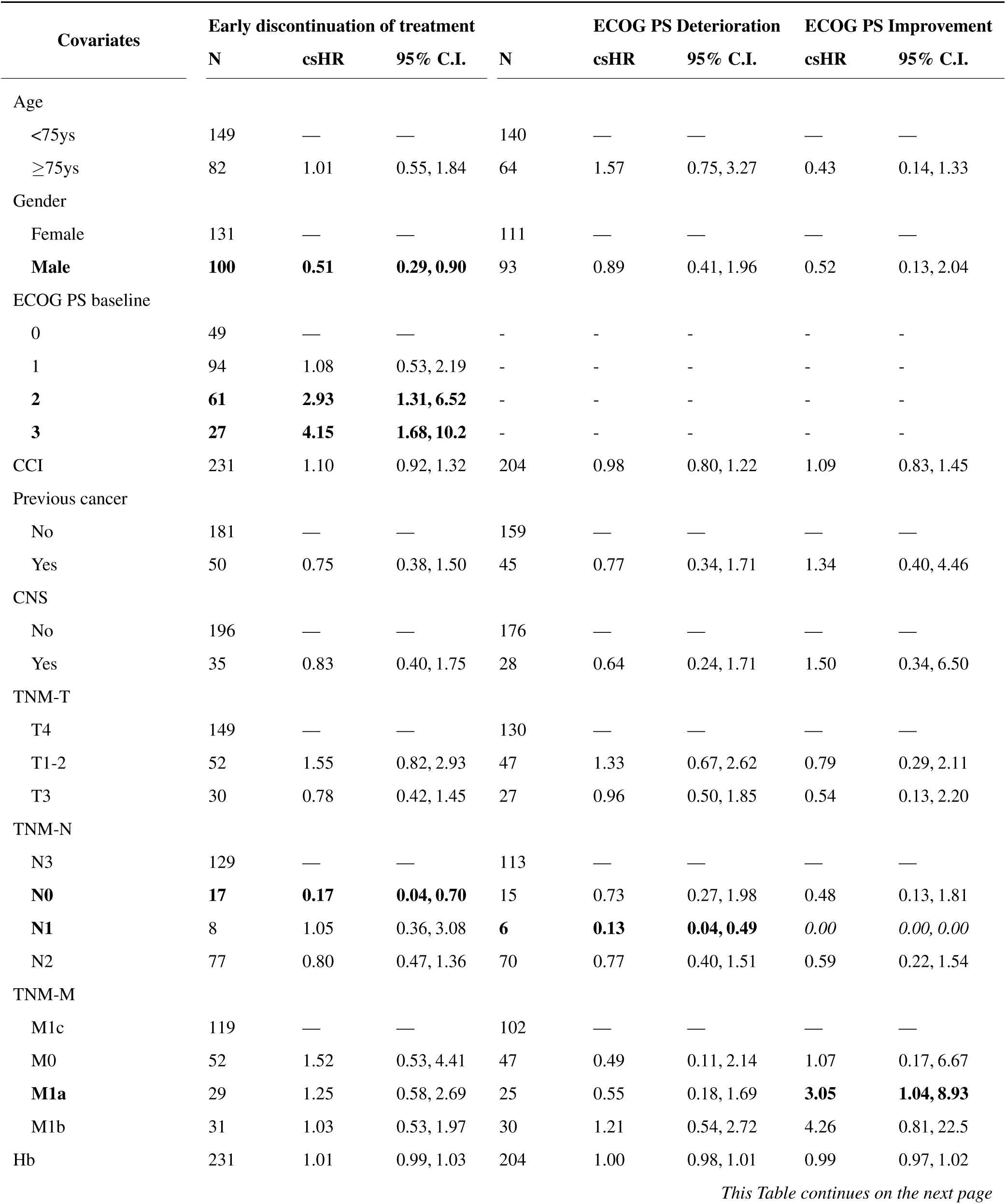
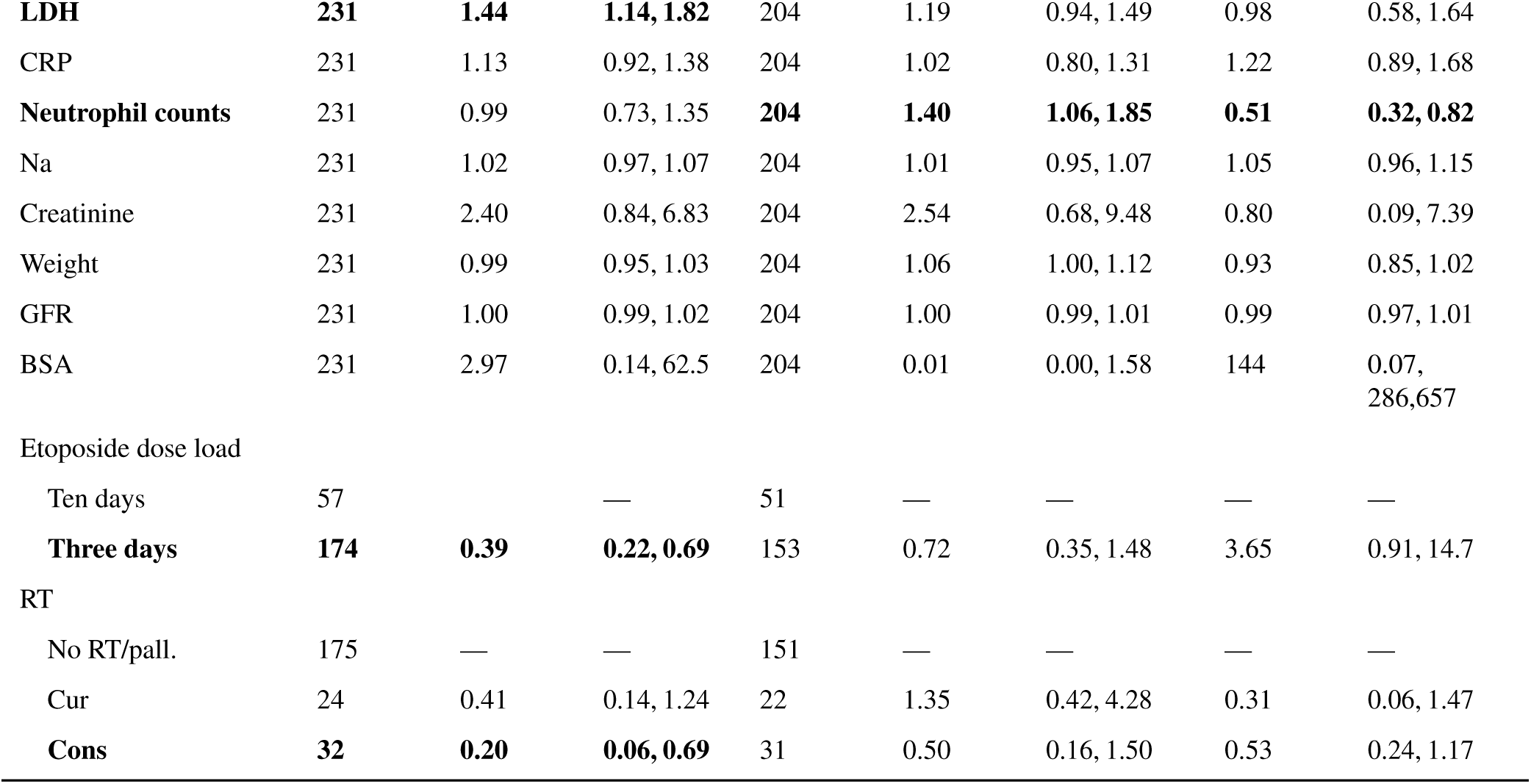
Competitive risk analysis results for the first line treatment platinum etoposide. csHR: cause specific Hazard Ratio, C.I.: confidence interval, ECOG PS: performance status, CCI: Charles Comorbidity Index, CNS: brain metastasis, LDH: lactate hydrogenasis, CRP: c-reactive protein, Na: sodium, Hb: hemoglobin, BSA: body surface area, GFR: estimated glomerular filtration rate, RT: radiotherapy, cur: curative, cons: consolidating, pall: palliative.

| Covariates | Early discontinuation of treatment |  |  | ECOG PS Deterioration |  |  | ECOG PS Improvement |  |
| --- | --- | --- | --- | --- | --- | --- | --- | --- |
|  | N | csHR | 95% C.I. | N | csHR | 95% C.I. | csHR | 95% C.I. |
| Age |  |  |  |  |  |  |  |  |
| <75ys | 149 | — | — | 140 | — | — | — | — |
| ≥75ys | 82 | 1.01 | 0.55, 1.84 | 64 | 1.57 | 0.75, 3.27 | 0.43 | 0.14, 1.33 |
| Gender |  |  |  |  |  |  |  |  |
| Female | 131 | — | — | 111 | — | — | — | — |
| **Male** | **100** | **0.51** | **0.29, 0.90** | 93 | 0.89 | 0.41, 1.96 | 0.52 | 0.13, 2.04 |
| ECOG PS baseline |  |  |  |  |  |  |  |  |
| 0 | 49 | — | — | - | - | - | - | - |
| 1 | 94 | 1.08 | 0.53, 2.19 | - | - | - | - | - |
| **2** | **61** | **2.93** | **1.31, 6.52** | - | - | - | - | - |
| **3** | **27** | **4.15** | **1.68, 10.2** | - | - | - | - | - |
| CCI | 231 | 1.10 | 0.92, 1.32 | 204 | 0.98 | 0.80, 1.22 | 1.09 | 0.83, 1.45 |
| Previous cancer |  |  |  |  |  |  |  |  |
| No | 181 | — | — | 159 | — | — | — | — |
| Yes | 50 | 0.75 | 0.38, 1.50 | 45 | 0.77 | 0.34, 1.71 | 1.34 | 0.40, 4.46 |
| CNS |  |  |  |  |  |  |  |  |
| No | 196 | — | — | 176 | — | — | — | — |
| Yes | 35 | 0.83 | 0.40, 1.75 | 28 | 0.64 | 0.24, 1.71 | 1.50 | 0.34, 6.50 |
| TNM-T |  |  |  |  |  |  |  |  |
| T4 | 149 | — | — | 130 | — | — | — | — |
| T1-2 | 52 | 1.55 | 0.82, 2.93 | 47 | 1.33 | 0.67, 2.62 | 0.79 | 0.29, 2.11 |
| T3 | 30 | 0.78 | 0.42, 1.45 | 27 | 0.96 | 0.50, 1.85 | 0.54 | 0.13, 2.20 |
| TNM-N |  |  |  |  |  |  |  |  |
| N3 | 129 | — | — | 113 | — | — | — | — |
| **N0** | **17** | **0.17** | **0.04, 0.70** | 15 | 0.73 | 0.27, 1.98 | 0.48 | 0.13, 1.81 |
| **N1** | **8** | **1.05** | **0.36, 3.08** | **6** | **0.13** | **0.04, 0.49** | *0.00* | *0.00, 0.00* |
| N2 | 77 | 0.80 | 0.47, 1.36 | 70 | 0.77 | 0.40, 1.51 | 0.59 | 0.22, 1.54 |
| TNM-M |  |  |  |  |  |  |  |  |
| M1c | 119 | — | — | 102 | — | — | — | — |
| M0 | 52 | 1.52 | 0.53, 4.41 | 47 | 0.49 | 0.11, 2.14 | 1.07 | 0.17, 6.67 |
| **M1a** | **29** | **1.25** | **0.58, 2.69** | **25** | **0.55** | **0.18, 1.69** | **3.05** | **1.04, 8.93** |
| M1b | 31 | 1.03 | 0.53, 1.97 | 30 | 1.21 | 0.54, 2.72 | 4.26 | 0.81, 22.5 |
| Hb | 231 | 1.01 | 0.99, 1.03 | 204 | 1.00 | 0.98, 1.01 | 0.99 | 0.97, 1.02 |
| **LDH** | **231** | **1.44** | **1.14, 1.82** | 204 | 1.19 | 0.94, 1.49 | 0.98 | 0.58, 1.64 |
| CRP | 231 | 1.13 | 0.92, 1.38 | 204 | 1.02 | 0.80, 1.31 | 1.22 | 0.89, 1.68 |
| **Neutrophil counts** | 231 | 0.99 | 0.73, 1.35 | **204** | **1.40** | **1.06, 1.85** | **0.51** | **0.32, 0.82** |
| Na | 231 | 1.02 | 0.97, 1.07 | 204 | 1.01 | 0.95, 1.07 | 1.05 | 0.96, 1.15 |
| Creatinine | 231 | 2.40 | 0.84, 6.83 | 204 | 2.54 | 0.68, 9.48 | 0.80 | 0.09, 7.39 |
| Weight | 231 | 0.99 | 0.95, 1.03 | 204 | 1.06 | 1.00, 1.12 | 0.93 | 0.85, 1.02 |
| GFR | 231 | 1.00 | 0.99, 1.02 | 204 | 1.00 | 0.99, 1.01 | 0.99 | 0.97, 1.01 |
| BSA | 231 | 2.97 | 0.14, 62.5 | 204 | 0.01 | 0.00, 1.58 | 144 | 0.07,<br>286,657 |
| Etoposide dose load |  |  |  |  |  |  |  |  |
| Ten days | 57 |  | — | 51 | — | — | — | — |
| **Three days** | **174** | **0.39** | **0.22, 0.69** | 153 | 0.72 | 0.35, 1.48 | 3.65 | 0.91, 14.7 |
| RT |  |  |  |  |  |  |  |  |
| No RT/pall. | 175 | — | — | 151 | — | — | — | — |
| Cur | 24 | 0.41 | 0.14, 1.24 | 22 | 1.35 | 0.42, 4.28 | 0.31 | 0.06, 1.47 |
| **Cons** | **32** | **0.20** | **0.06, 0.69** | 31 | 0.50 | 0.16, 1.50 | 0.53 | 0.24, 1.17 |

The results show a strong association between higher etoposide dose levels and the subsequent occurrence of AEs (OR: 5.97 [1.41, 30.5]), specifically neutropenia (OR: 3.55 [1.03, 13.3]). Early discontinuation of therapy was associated to higher LDH levels (csHR 1.44 [1.14, 1.82]), ECOG PS 2 (csHR: 2.93 [1.31, 6.52], ECOG PS 3 (csHR 4.15 [1.68, 10.2]).

Males who had a complete first line treated course had a longer PFS. (csHR: 0.51 [0.29, 0.90]). As did subgroups of patients with TNM-M0 and those receiving consolidating radiotherapy (being mostly TNM-IV, n=29) (Table 3). Similarly, patients with M1a status regardless of T and N stage were associated with ECOG PS improvement (Table 3).

#### Second line therapy PE-IP

Results of the multivariate analysis for the second-line treatment are reported in Supplementary Tables S4 and S5. Due to the large confidence intervals of the ORs, the ORs are reported on a log scale.

A higher dose of platinum was associated with a higher incidence of AEs and a higher risk of neutropenia. Patients receiving IP experienced fewer AEs. Male patients were less likely to experience AEs and neutropenia. However, male patients were associated with worse PFS (HR: 1.62 [1.10, 2.38]) due to early discontinuation of the second line regardless PE or IP (n=44). Higher sodium levels were instead associated with lower risk of neutropenia and ECOG PS deterioration. Patients with TNM-M M1a had a higher probability of ECOG PS deterioration and more AEs. CNS metastasis was linked to increased risk for ECOG PS deterioration and early discontinuation of therapy.

#### High-risk subpopulation analysis: to treat or not to treat?

In the Supplementary Tables S6-11 we report the results of the multivariate analysis for the high-risk subgroups. Table 4 summarises the significant covariates for each subgroup. Similarly to the second line therapy analysis, the ORs were reported in log scale due to the wide confidence intervals.

**Table 4.** Summary of statistically significant covariates in the multivariate analysis for high-risk subgroups. (+): OR and csHR with a direct effect on the studied outcome, (-): OR and csHR with an inverse effect on the studied outcome, AEs: adverse effects, GT: grade of toxicity, ECOG PS: ECOG performance status, CCI: Charles Comorbidity Index, LDH: lactate hydrogenasis, CRP: c-reactive protein, BSA: body surface area.

| High risk Subgroup | AEs occurrence | Neutropenia (GT) | ECOG PS deterioration | ECOG PS improvement | Early Discontinuation |
| --- | --- | --- | --- | --- | --- |
| TNM-IVB | Dose etoposide (+)<br>Dose*interval etoposide (+)<br>Albumin (-)<br>Neutrophil counts (-) | Dose*interval etoposide(+)<br>BSA (-) | Neutrophil counts(+) | Neutrophil counts(-)<br>LDH(+) | LDH(+)<br>Creatinine (+) |
| ECOG PS 2-3 | Dose*interval platinum (+) | Dose*interval platinum (+) | Neutrophil counts (+) | Neutrophil counts (-)<br>LDH (+) | Etoposide 10-days regime (+)<br>TNM-N0 (-)<br>TNM-M0 (+)<br>HB (+)<br>LDH (+)<br>CRP (+)<br>CCI (+) |
| AGE ≥ 75 | LDH (+)<br>Albumin (-)<br>Neutrophil counts (-)<br>Male (-)<br>ECOG 3 (-)<br>TNM-T3 (-)<br>TNM-N2 (+) | Dose*interval etoposide (+)<br>weight (-)<br>Male (-)<br>TNM-N0 (+)<br>TNM-M1a (+) | TNM-N2 (-) | | ECOG 2 (+)<br>TNM-N0 (-)<br>LDH (+) |

Results observed in the subgroup analyses that aligned with the ones of the overall cohort were the association of LDH and ECOG PS 2-3 with early discontinuation of therapy (all subgroups), dose etoposide and increased risk of AEs and neutropenia (IVB and age *≥* 75ys), lower AE risk for male patients (age *≥* 75ys). For patients with ECOG PS 2-3 higher dose of platinum was associated with an increased risk of AEs and neutropenia.

The comparison with patients that did not receive chemotherapy showed that, even though AEs were associated with longer survival, the hazard ratios of patients with ECOG PS 2-3 or age *≥* 75ys were closer to the non-treated subgroup rather than to the patients who experienced a good response to treatment (Supplementary Table S12). We observed that when further stratifying the high-risk subgroups using the significant variables reported in Table 4, the subgroups showed similar survival to the same group of patients that did not receive treatment. For example, Figure 4 shows the Kaplan Meier curves of the comparative analysis for patients with ECOG PS 2-3 and age *≥* 75 years.

**Figure 4.**
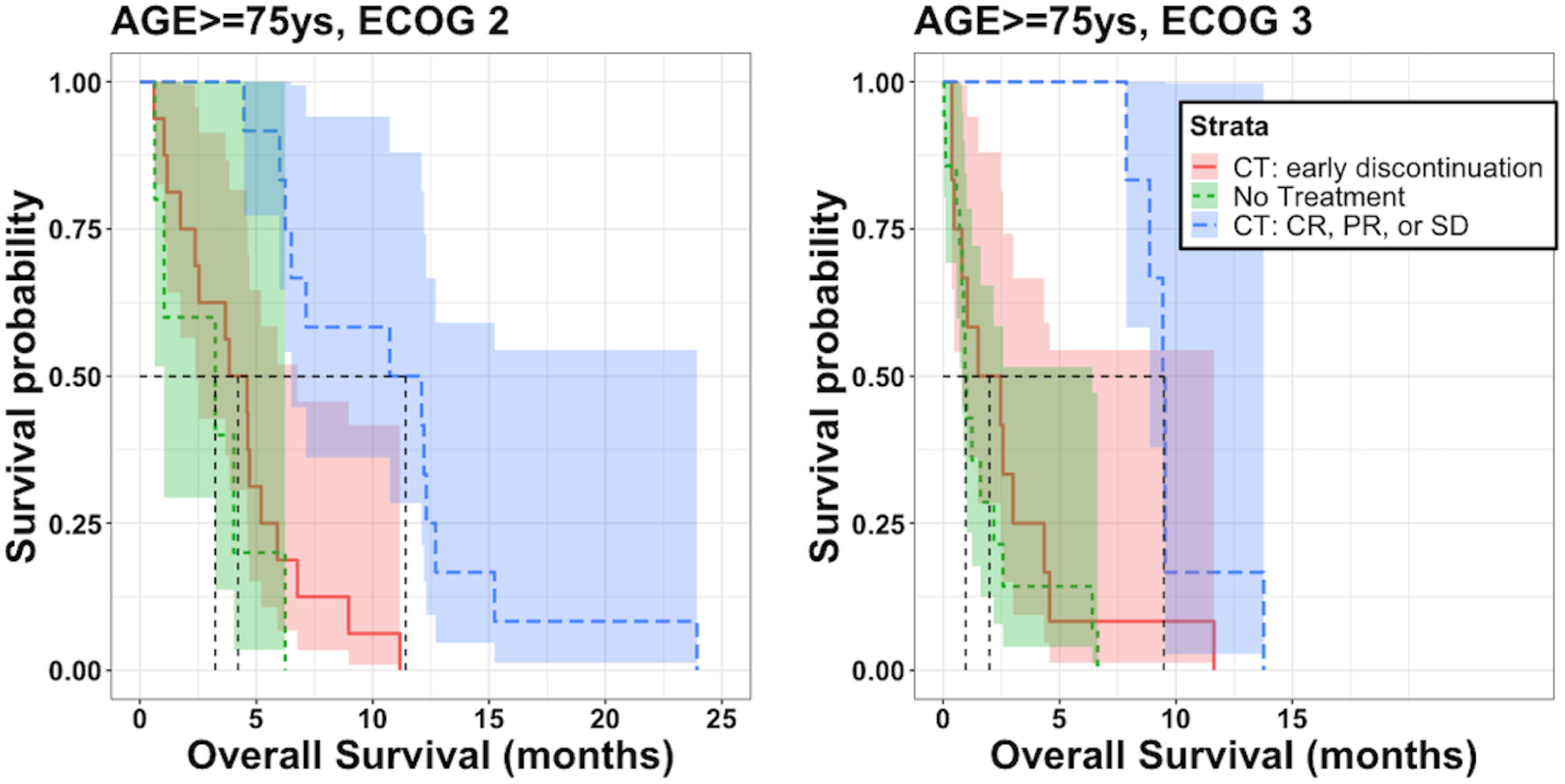
Kaplan Meier curves of patients older than 75 years (yrs) with ECOG performance status (PS) 2-3 stratified by good response to first line chemotherapy, unplanned stop of treatment, and patients that did not received chemotherapy. CT: chemotherapy, CR: complete response, PR: partial response, SD: stable disease.

## Discussion

In this study, we analysed a longitudinal real-world cohort of SCLC patients and their treatment pathways. The analysis allowed the study of the relationships between treatment decisions, phases of progression of disease, occurrence of AEs, and overall survival. To the best of our knowledge, this is one of the most detailed longitudinal datasets of SCLC treatment decisions.

The improved prognosis of patients that experienced neutropenia align with previous findings^40,41^. The analysis allowed the investigation of treatment-related adverse events in relation to efficacy of therapy, or deterioration of the patient conditions (i.e., Figure 2). We identified significant ORs relating to the dose of etoposide and increased occurrence of AEs. The analysis showed a similar OS for patients receiving four doses during first line therapy, regardless of if the patient had dose adjustment/reduction or not (Figure 3). Worse prognosis was observed when there was a deterioration in ECOG PS or early discontinuation of treatment.

SCLC treatment effects vary widely between individuals. This could suggest the need for individualised dosing^10,11,42^ tailored to balance improved survival and reduced incidence of AEs in the first line treatment. Indeed, etoposide dose calculation is based on BSA that has been shown to present such limitations^43^. Similar analyses could be done for the second line. In fact, only a few patients received the full expected dose without dose reduction, and the platinum dose and PE treatment were strongly associated with AEs.

ECOG PS 2-3 and elevated LDH serum levels are known to be prognostic factors of worse OS in SCLC^8,44^. Our results added a new insight by showing their role in predicting early discontinuation of treatment, which was associated with significantly poorer OS. Another finding was the high probability of a favourable response to first-line therapy in male patients, along with their lower likelihood of experiencing AEs (Table 3-4 and Supplementary Tables S4-5). Moreover, when stratifying older patients by sex, women who developed neutropenia exhibited survival rates comparable to those in the same subgroup who did not receive chemotherapy. A possible explanation is that females, a group that historically has been underrepresented in clinical trials^9,45,46^, may be overtreated, which increases the risk of AEs, negatively impacting their overall prognosis. In addition, the second-line therapy worse PFS and high frequent early discontinuation for male patients might suggest the need of further studies regarding different therapy responses for males and females.

The high-risk subgroup analysis allowed detection of key groups of patients, such as elderly patients with poor ECOG PS, where an eventual poor response to therapy is associated with similar survival as compared to patients not receiving chemotherapy. This finding is clinically relevant, as it highlights the importance of balancing benefit and risk of treating such fragile patients and may help clinicians avoid overtreating this vulnerable population. These outcomes underline the potential role that real-world data may fill in providing evidence on treatment in groups of patients that are underrepresented in clinical trials^9^.

The design of the analytic pipeline relied on the consideration that the data should be analysed using models capable of capturing the complexity of the patient treatment pathways without sacrificing interpretability (such as, black box models^23^). We believe that this is an important consideration when approaching real-world data. In fact, the discussions with the clinicians on the obtained results allowed the assessment of the validity of the results, the detection of potential inherent bias, and non-observable effects. For example, it was observed that the 10-day dosing regimen of etoposide resulted in poorer survival. At the time of the present analysis, this dosing regimen had, however, already been abandoned due to toxicity caused by the limited time for patients to recover between the doses (Tables 2-3).

The study had several limitations. The reason for missing values was not at random for some of the variables, thus introducing some biases on the results. Particularly for baseline neutrophil counts, where missing observations were high across all treatment lines. Further, these variables were available only at the start of the treatment line, meaning that we were not able to analyse their time-profiles (i.e., neutrophil counts over time between doses). Moreover, it should be stated that AEs are not always described in medical notes, thus introducing investigator bias in the collected data.

Another limitation was the sample size of the high-risk subgroups and patients receiving the second line treatment, resulting in higher estimates of variability of the outcomes as compared to the first line treatment cohort. In detail, the odds ratios of the mixed effects models resulted in wide confidence intervals. The reason of this high variability in outcomes could be due to the fact that these small cohorts were also associated with fewer longitudinal observations per patient (the number of administrated doses). Moreover, patients receiving more than two lines were not included in the multivariate analysis, as this was only observed in few instances.

The median age of our population was 72 years, yet we chose 75 years to better isolate a more vulnerable subset of patients by moving further from the median. This aligns with existing studies that identify this age group as particularly fragile, often requiring specialized considerations in treatment planning^47^. Future studies exploring different criteria and strategies to separate elderly patients would be required.

This was a single-centre study, and expanding the analysis by including other SCLC cohorts from other centres would be beneficial. A comparison of the dose adaptation and the follow-up processes in clinical trials would be of high importance^9^. A future aspect to explore would be data collection, designed similarly to the study presented here, in larger initiatives such as cancer registries^48,49^.

This study could inform the design of future works, such as multi-state models of patient survival in function of different therapies and events^50,51^, or model-informed precision dosing models to explore individualised treatment and dosing to reduce the risk of AEs (e.g., neutropenia) without compromising the efficacy of the treatment in terms of survival outcomes^52,53^. A continued fruitful collaboration between clinical and technical experts when analysing complex data and contextualising the clinical processes would enhance the impact of real-world data in clinical practice and clinical drug development.

## Author contributions

L.M. and A.D. wrote the manuscript. L. M., A.S.D., A.D., L.D.P., S.T., J.R., R.L., and S.M. designed the research. L.M., A. D., L.D.P. and A.S.D. performed the research. L.M., A.D., A.S.D., and L.D.P. analysed the data.

## Data availability

The data that support the findings of this study are not openly available due to reasons of sensitivity and are available from the authors upon reasonable request. Data are located in a controlled access data storage at Karolinska University Hospital.

