## Supplementary Figures for "Real-World Evidence on Dose-Reduction and Treatment Outcomes in Small Cell Lung Cancer; A Bayesian mixed effects and Competitive Risk Approach"

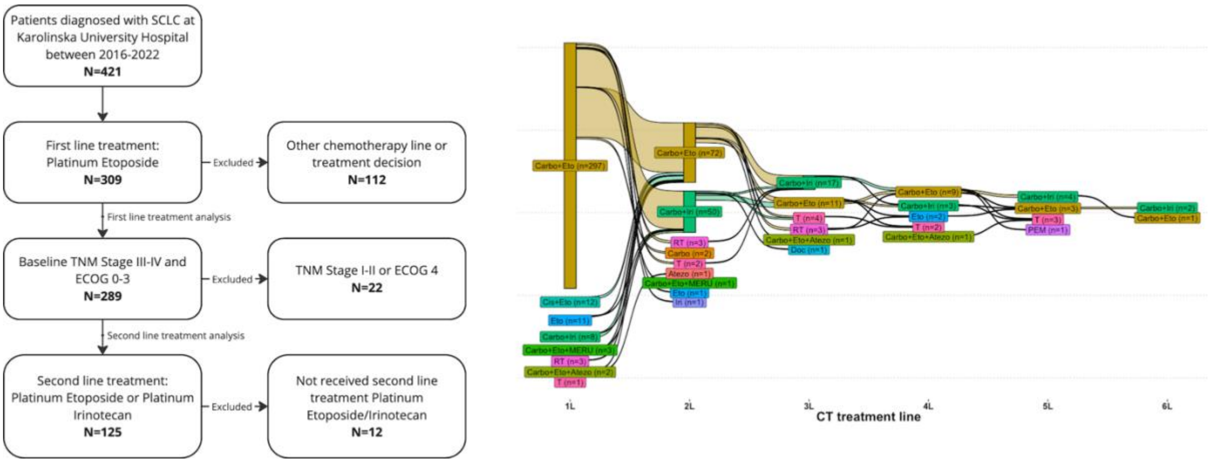


Figure S1. Sankey Flow of the subsequent chemotherapy treatment lines.

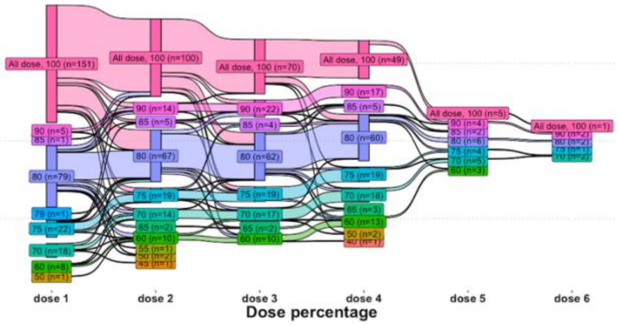


Figure S2. Sankey flow or the subsequent doses percentages for patients receiving first line platinum etoposide.

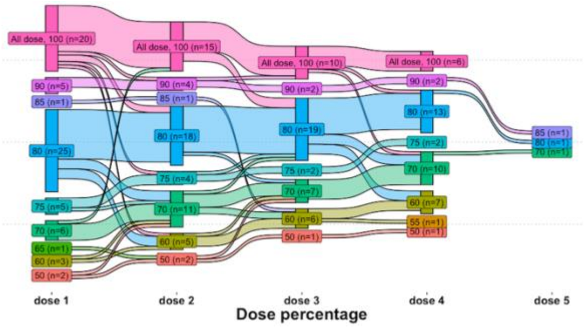


Figure S3. Sankey flow or the subsequent doses percentages for patients receiving second line platinum etoposide after first line platinum etoposide.

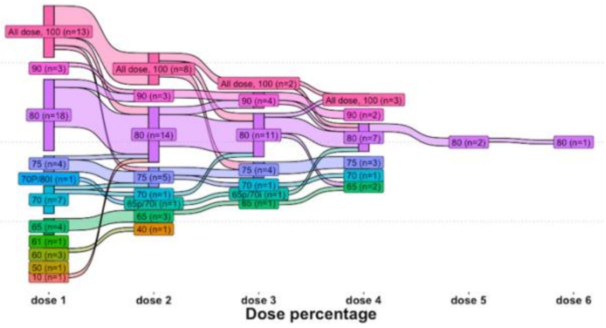


Figure S4. Sankey flow or the subsequent doses percentages for patients receiving second line platinum irinotecan after first line platinum etoposide.

### Supplementary Tables

Supplementary Tables S1-S12 are reported in Supplementary_Tables.xlsx file.
